# Identifying proteomic risk factors for overall, aggressive and early onset prostate cancer using Mendelian randomization and tumor spatial transcriptomics

**DOI:** 10.1101/2023.09.21.23295864

**Authors:** Trishna A Desai, Åsa K Hedman, Marios Dimitriou, Mine Koprulu, Sandy Figiel, Wencheng Yin, Mattias Johansson, Eleanor L Watts, Joshua R Atkins, Aleksandr V Sokolov, Helgi B Schiöth, Marc J Gunter, Konstantinos K Tsilidis, Richard M Martin, Maik Pietzner, Claudia Langenberg, Ian G Mills, Alastair D Lamb, Anders Mälarstig, Tim J Key, The PRACTICAL Consortium, Ruth C Travis, Karl Smith-Byrne

## Abstract

**Background:** Understanding the role of circulating proteins in prostate cancer risk can reveal key biological pathways and identify novel targets for cancer prevention.

**Methods:** We investigated the association of 2,002 genetically predicted circulating protein levels with risk of prostate cancer overall, and of aggressive and early onset disease, using *cis-*pQTL Mendelian randomization (MR) and colocalization. Findings for proteins with support from both MR, after correction for multiple-testing, and colocalization were replicated using two independent cancer GWAS, one of European and one of African ancestry. Proteins with evidence of prostate-specific tissue expression were additionally investigated using spatial transcriptomic data in prostate tumor tissue to assess their role in tumor aggressiveness. Finally, we mapped risk proteins to drug and ongoing clinical trials targets.

**Results:** We identified 20 proteins genetically linked to prostate cancer risk (14 for overall [8 specific], 7 for aggressive [3 specific], and 8 for early onset disease [2 specific]), of which a majority were novel and replicated. Among these were proteins associated with aggressive disease, such as PPA2 [Odds Ratio (OR) per 1 SD increment = 2.13, 95% CI: 1.54-2.93], PYY [OR = 1.87, 95% CI: 1.43-2.44] and PRSS3 [OR = 0.80, 95% CI: 0.73-0.89], and those associated with early onset disease, including EHPB1 [OR = 2.89, 95% CI: 1.99-4.21], POGLUT3 [OR = 0.76, 95% CI: 0.67-0.86] and TPM3 [OR = 0.47, 95% CI: 0.34-0.64]. We confirm an inverse association of MSMB with prostate cancer overall [OR = 0.81, 95% CI: 0.80-0.82], and also find an inverse association with both aggressive [OR = 0.84, 95% CI: 0.82-0.86] and early onset disease [OR = 0.71, 95% CI: 0.68-0.74]. Using spatial transcriptomics data, we identified MSMB as the genome-wide top-most predictive gene to distinguish benign regions from high grade cancer regions that had five-fold lower MSMB expression. Additionally, ten proteins that were associated with prostate cancer risk mapped to existing therapeutic interventions.

**Conclusion:** Our findings emphasize the importance of proteomics for improving our understanding of prostate cancer etiology and of opportunities for novel therapeutic interventions. Additionally, we demonstrate the added benefit of in-depth functional analyses to triangulate the role of risk proteins in the clinical aggressiveness of prostate tumors. Using these integrated methods, we identify a subset of risk proteins associated with aggressive and early onset disease as priorities for investigation for the future prevention and treatment of prostate cancer.

## Introduction

Prostate cancer is a heterogeneous disease with a high survival rate for those diagnosed with indolent or low-stage disease, but a less than 50% 5-year survival rate for those diagnosed with aggressive or metastatic cancer.^1^ The proportion of these clinically aggressive cases is higher among men younger than 55 years (early onset disease), which contributes to premature death among these men.^2,3^ However, few risk factors for prostate cancer have been established. These include: advanced age, African ancestry, family history of the disease, circulating levels of insulin-like growth factor I and microseminoprotein-beta (MSMB), with little evidence for successful strategies for prevention. ^4–7^

Recent advances in multiplexed and high throughput platforms as well as the widespread availability of genotypic arrays have identified genetic variants that determine circulating levels of thousands of circulating proteins, known as protein-quantitative trait loci (pQTL). PQTL, in particular those lying in or near a protein’s cognate gene (referred to as *cis*-pQTL), can be leveraged to identify candidate etiological proteins for cancer risk through Mendelian randomization (MR) analyses, an approach that can limit the impact of reverse causality.^8,9^ MR can also be complemented with colocalization analyses to further exclude confounding by linkage disequilibrium (LD).^10^ Candidate etiological proteins for cancer risk identified using these methods can provide a valuable starting point for further analyses using more resource-intense methods, such as spatial transcriptomics, where their functional importance at the tissue level can be directly interrogated to triangulate their role in etiology.^11,12^

Using an integrated *cis-*pQTL MR and colocalization pipeline, we analyzed the associations of 2,002 unique proteins with overall, aggressive, and early onset prostate cancer and replicated and mapped those with significant findings to drug targets. Additionally, we investigated the spatial distribution and gene expression profiles of a subset of these proteins in prostate tumor tissue using spatial transcriptomics. In doing so, we demonstrate the value of protein MR and colocalization analyses to identify proteins that may have a causal role in the tumor aggressiveness.

## Methods

### Overall study design

We extracted *cis* genetic instruments for circulating protein levels from publicly available datasets, and harmonized these *cis*-pQTL with the GWAS results from an international prostate cancer consortium, including aggressive and early onset subtypes (Supplementary Figure 1). We subsequently estimated risk associations for protein levels using *cis*-pQTL MR against each of these three prostate cancer endpoints. All associations passing a multiple testing threshold in MR analyses were then followed up with colocalization analyses. Where data were available, we performed replication analyses in an external prostate cancer GWAS in European and African ancestry (using African ancestry specific *cis*-pQTLs – see below) populations for proteins with evidence from MR and colocalization analyses. For proteins identified as risk factors for prostate cancer with evidence of specific expression in the prostate tissue, we additionally performed analyses using spatial transcriptomics to gain insights into the spatial distribution and gene expression patterns of these proteins in prostate tumor samples. Finally, we conducted an exploratory analysis restricting to *cis-*pQTL whose cognate genes are established drug targets.

### Identification of cis-pQTL

Genetic instruments for *cis*-pQTL were extracted from 4 publicly available protein GWAS at p < 5 x 10^−8^ and clumped at R^2^=0.01 within their originating panel (instruments presented in Supplementary Table 1).^13–16^ *Cis*-instruments were defined in the first instance as those that were genome-wide significant (p< 5 x 10^−8^) within 1 Mb of the transcription start side of the measured protein encoded gene, or as the sentinel *cis*-pQTL for the measured protein depending on data availability. We additionally gathered data on *cis-*pQTL from published GWAS present on the OpenGWAS platform using a relaxed p-value threshold of 5 x 10^-5^ due to the high biological plausibility of identifying *cis-*pQTL at or near a protein’s cognate gene.^17,18^ Specifically, we extracted unreported *cis*-pQTL from the genomic region 1 megabase up and downstream of the cognate gene for a given protein GWAS (Supplementary Table 1). We subsequently extracted all instruments where no *cis*-pQTL was present at p < 5 x 10^-8^ but at least one *cis*-pQTL was present at p < 5 x 10^-05^.

All instruments were mapped to Uniprot IDs, and *cis-*pQTL with weak instrument strength at F_stat_ < 10 [β^2^/σ^2^] were excluded from the study. For *cis*-pQTL that were not present in the cancer outcome data, SNP proxies were selected at r^2^ where r^2^ > 0.8 in 1000 genomes CEU population with the index *cis-*pQTL. In total, 2,002 unique plasma proteins that fit these criteria were included in analyses.

### Cancer outcome data

Genetic associations for overall, aggressive, and early onset prostate cancer were obtained from the Prostate Cancer Association Group to Investigate Cancer Associated Alterations in the Genome (PRACTICAL) consortium (Supplementary material).^19^ Full study characteristics have been described previously, but briefly, summary statistics for SNP associations with prostate cancer and subtypes were generated from the PRACTICAL consortium using 85,554 overall prostate cancer cases and 91,972 controls (database of Genotypes and Phenotypes [dbGaP] project #31553), 15,167 aggressive PC cases and 58,308 controls, and 6,988 cases of early onset disease and 44,256 controls, all of European ancestry.^20,21^ Aggressive prostate cancer is defined in PRACTICAL as cases having metastatic disease or Gleason score >=8 or PSA >100 ng/mL or prostate cancer death. Early onset PC cases are defined as those diagnosed before the age of 55 years. Genotype information was imputed for samples using the 2014 release of the 1000 Genomes Project as a reference panel.

### Two-sample Mendelian randomization

*Cis-*pQTL data were harmonized to each cancer outcome by rsID and oriented to the protein-increasing allele. Two-sample Mendelian randomization (MR) was subsequently performed for each *cis*-pQTL on risk of overall, aggressive, and early onset prostate cancer using the Wald-ratio method (*β_cancer/_β_protein_*). Resulting associations where the *p*_Wald_ passed a Bonferroni-corrected threshold of significance based on the total number of unique proteins assessed for each of the three prostate cancer outcomes were taken forward in analyses (*p*_Wald_ < 0.05/N_Proteins analyzed per cancer outcome_).^22^ Multiple independent [*r*^2^ < 0.01] *cis*-pQTL that proxied the same protein and were both associated after correction for multiple testing and that colocalised with the same prostate cancer outcome were combined using the inverse-variance weighted method (IVW). Odds ratio estimates are scaled per standard deviation increment in relative and normalized circulating protein concentrations.

### Colocalization

Colocalization was performed to assess the probability that the protein and cancer instruments share a causal variant, fulfilling an important instrumental variable assumption of MR.^9,10^ Specifically, single and conditional iterative colocalization analysis were performed for all *cis-*pQTL MR results that passed a Bonferroni correction for multiple testing based on the number of unique proteins in the study (p < 0.05/ N _Proteins_), using all variants within a 75kb region up- and downstream from the index *cis-*pQTL to assess confounding by linkage disequilibrium.^10,23^ To mitigate the chance of false-positive findings, we selected priors of P1: 1 x 10^-3^, P2: 1 x 10^-4^, and P12: 1 x 10 ^-5^, which roughly equate to a 0.1% prior belief in colocalization (PP4).^24^ We defined a threshold PP4 in support of a shared association for a protein and cancer signal at 0.70 to take proteins forward for subsequent analysis and the highest PP4 of any method of colocalization was recorded to assess confidence in the shared association for each SNP assessed.

### Replication of robust proteins in European and African ancestry populations

We conducted a replication analysis of *cis*-pQTL MR associations passing multiple testing correction and that colocalized (referred to as *robust* proteins) using an external GWAS in a European ancestry population of overall prostate cancer risk. GWAS summary statistics represented a meta-analysis in FinnGen r9 and the UK Biobank (20,907 cases & 289,710 controls).^25,26^ Additionally, where possible, we performed replication analyses using *cis-*pQTL identified in an African ancestry protein GWAS in the Atherosclerosis Risk in Communities study (4,657 proteins in 467 African-ancestry participants) and a GWAS of overall prostate cancer among African-ancestry populations obtained from dbGaP (project #31553) containing data from the AAPC GWAS, Ghana Prostate Study, ProHealth Kaiser GWAS, and ELLIPSE OncoArray (10,368 cases and 10,986 controls).^21,27^ GWAS for aggressive and early onset prostate cancer were unavailable to use as a replicate sample in either ancestry population. We considered a directionally concordant risk estimates and Wald ratio p < 0.05 using external data to indicate replication. No sample overlap was present between samples used to generate protein associations and used to conduct replication analyses.

### Drug target pQTL analyses

We restricted our MR results to those *cis*-pQTL that share a cognate gene that is an established drug target by reference to the DrugBank, Therapeutic Target Database, Pharos consortium, ClinicalTrials.gov or expert curation.^28–30^ As above, we defined *robust* associations as Wald p< 0.05/N_Proteins_, where N_Proteins_ is the number of unique proteins analyzed for a given cancer outcome that were identified as the cognate gene of a pharmaceutical target and PP4 > 0.7. Additionally, all proteins identified in overall and drug target analyses were queried the Cortellis database (https://www.cortellis.com) to assess the highest current level of clinical development stage.

Statistical analysis was performed in R version 4.1 and all tests of significance were two-sided, where *P* values <0.05 were considered statistically significant. MR analyses were performed using the *TwoSampleMR* R package and colocalization analyses were performed using the *coloc* R package.^17,23^

### Gene expression analysis using spatial transcriptomics

Spatial transcriptomics provides a spatial map of gene expression within the target tissue. This spatial information can be used to investigate the variation in gene expression by healthy tissue and tumor type intratumorally, and as a result, it can provide valuable insights into tumorigenesis and inform causal inference in this molecularly heterogeneous disease.^31,32^ Spatial transcriptomic analysis was performed for those proteins passing multiple testing correction and that colocalized and also showed high expression in the prostate epithelium.^33^ Data for spatial transcriptomics were obtained from our previously published dataset derived from radical prostatectomy tissue taken from a patient with multifocal prostate cancer.^34^ Our analysis focused on eight distinct tissue sections, which collectively comprised 32,156 spots, some of which contained regions of cancer as well as histo-pathologically benign prostate tissue, and some of which did not contain cancer. To ensure data quality, samples with less than 500 Unique Molecular Identifier (UMI) counts were excluded from the analysis. The initial fastq files were processed using the 10x Visium Spaceranger software, enabling the conversion of the files into gene expression data. Subsequently, the data underwent SCTransform normalization and variance reduction procedures. A consensus pathology approach was employed involving two pathologists who independently annotated each spatial transcriptomics spot, with the aim to include those that predominantly contained epithelial cells, which comprised approximately 1-15 cells. Violin plots were generated using Graphpad Prism (version 10).

### Iterative random forest network using spatial transcriptomics

We used the iterative random forest (iRF) method to investigate gene interactions.^35^ With this method, we randomly selected genes and constructed random forests with other genes as branches to identify the most robust gene expression network. The analysis specifically focused on comparing the gene interactions between benign and Gleason grade group 4 histology status. The criteria we used to select credible random forest model was stability > 0.8 and precision > 0.8. The resulting gene network was visualized using Gephi 0.99.

## Results

We investigated the associations of 2,002 unique proteins using 4,592 *cis*-pQTL that harmonized with the GWAS summary statistics for at least one of overall (1,999 proteins; 4,582 *cis*-pQTL), aggressive (1,986 proteins; 4,543 *cis*-pQTL), or early onset prostate cancer (1,984 proteins; 4,534 *cis*-pQTL) (Figure 1). From these analyses we identified 20 proteins that were associated, after correction for multiple testing, with at least one of overall (14 proteins), aggressive (7 proteins), or early-onset (8 proteins) prostate cancer and with support from colocalization analyses (Figure 2, Table 1). Of the 20 proteins associated with any prostate cancer outcome, several showed robust associations in only one outcome, including seven that appeared specific to overall prostate cancer (5NTC, CREBL1, INFA14, ISLR2, MMP7, SERPINA1, TNSFRS10B), three that appeared specific to aggressive disease (C4A, C2, TNFRSF6B), and two that appeared specific to early onset disease (SERPINA3, PYY).

**Figure 1.**
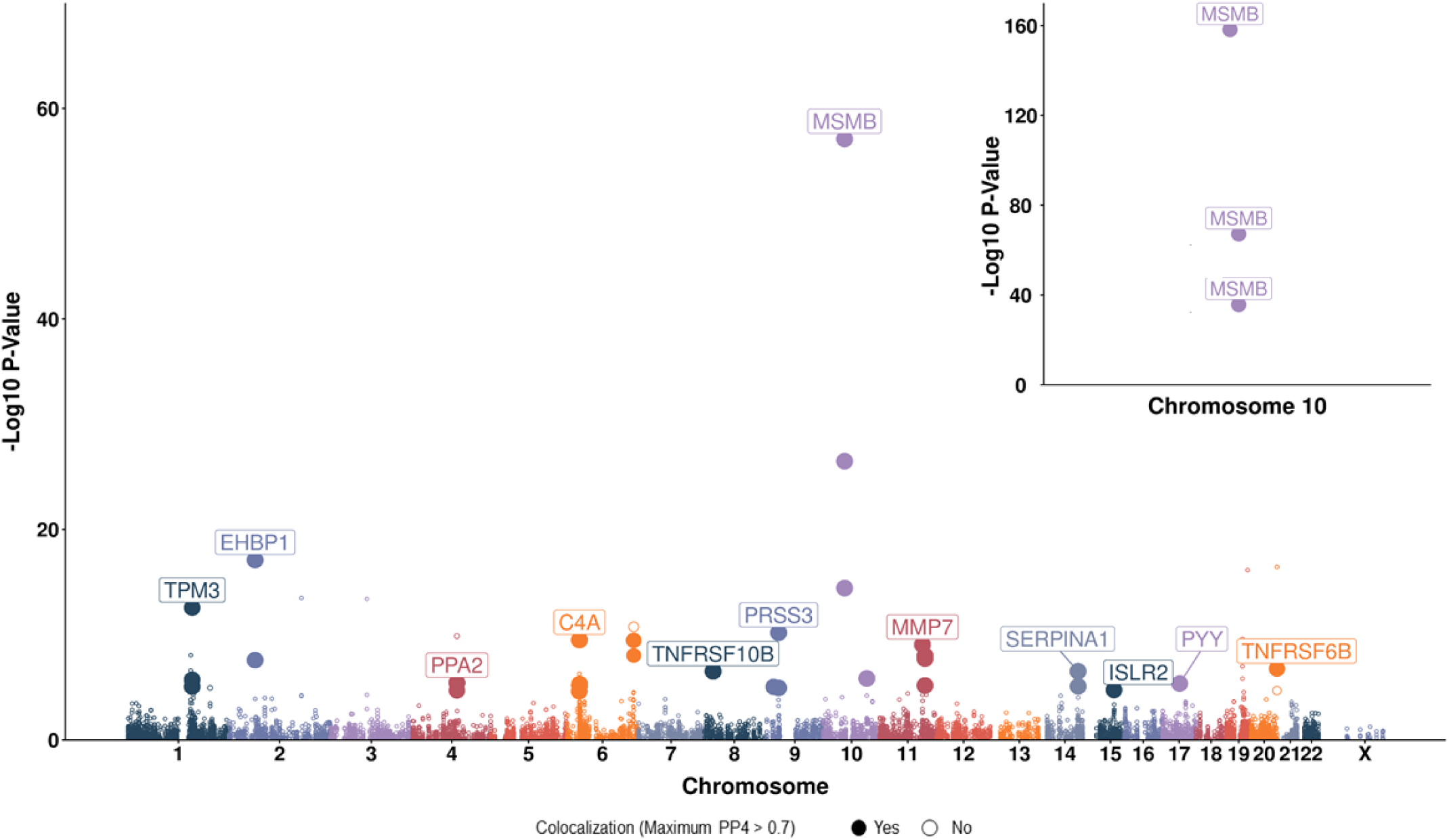
Association of genetically predicted protein concentrations with prostate cancer risk presented as a Manhattan plot where position is given by *cis-*pQTL coordinate (chromosome and base-pair position) labelled with their association with cancer risk and the highest colocalization probability from single or conditional iterative methods (PP4). Points highlighted as filled-in are those with evidence of a shared causal locus (PP4 > 0.7) with point size reflecting PP4 magnitude, which can vary between 0 and 1. Risk associations with MR *p* > Bonferroni correction threshold were not subject to colocalization analyses. The strongest protein-cancer association per chromosome is labelled and a zoomed-in plot for MSMB (rs10993994) on chromosome 10 is shown in the upper right-hand corner.

**Figure 2.**
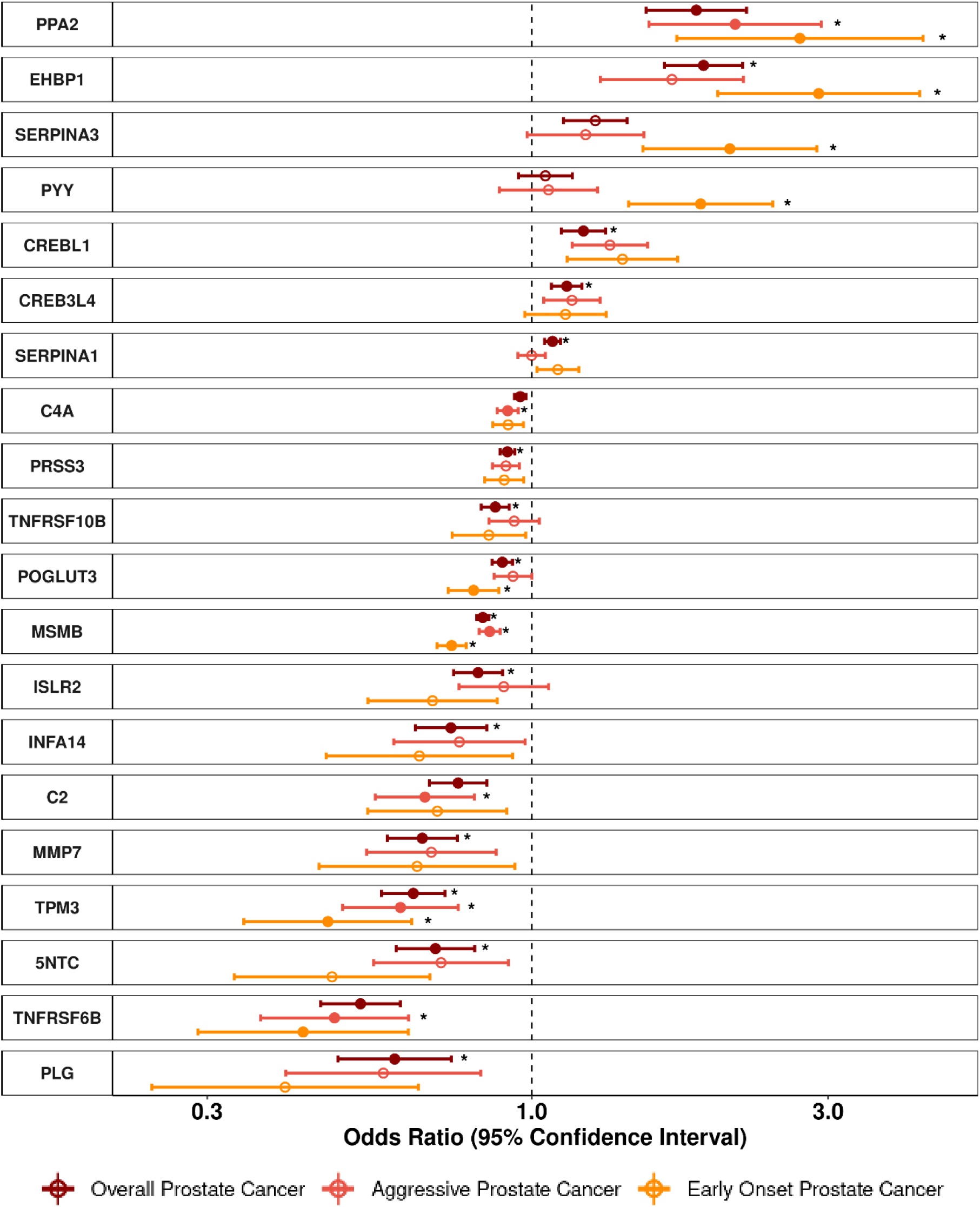
Odds ratios (95% confidence intervals) for genetically predicted protein levels and prostate cancer risk (for proteins with p< Bonferroni threshold based on 0.05/number of proteins analyzed). Odds ratio estimates are scaled per standard deviation increment in genetically predicted relative circulating protein concentrations. Filled circles represent Bonferroni-significant associations and asterisks indicate evidence for colocalization (PP4 > 0.70).

**Table 1.**
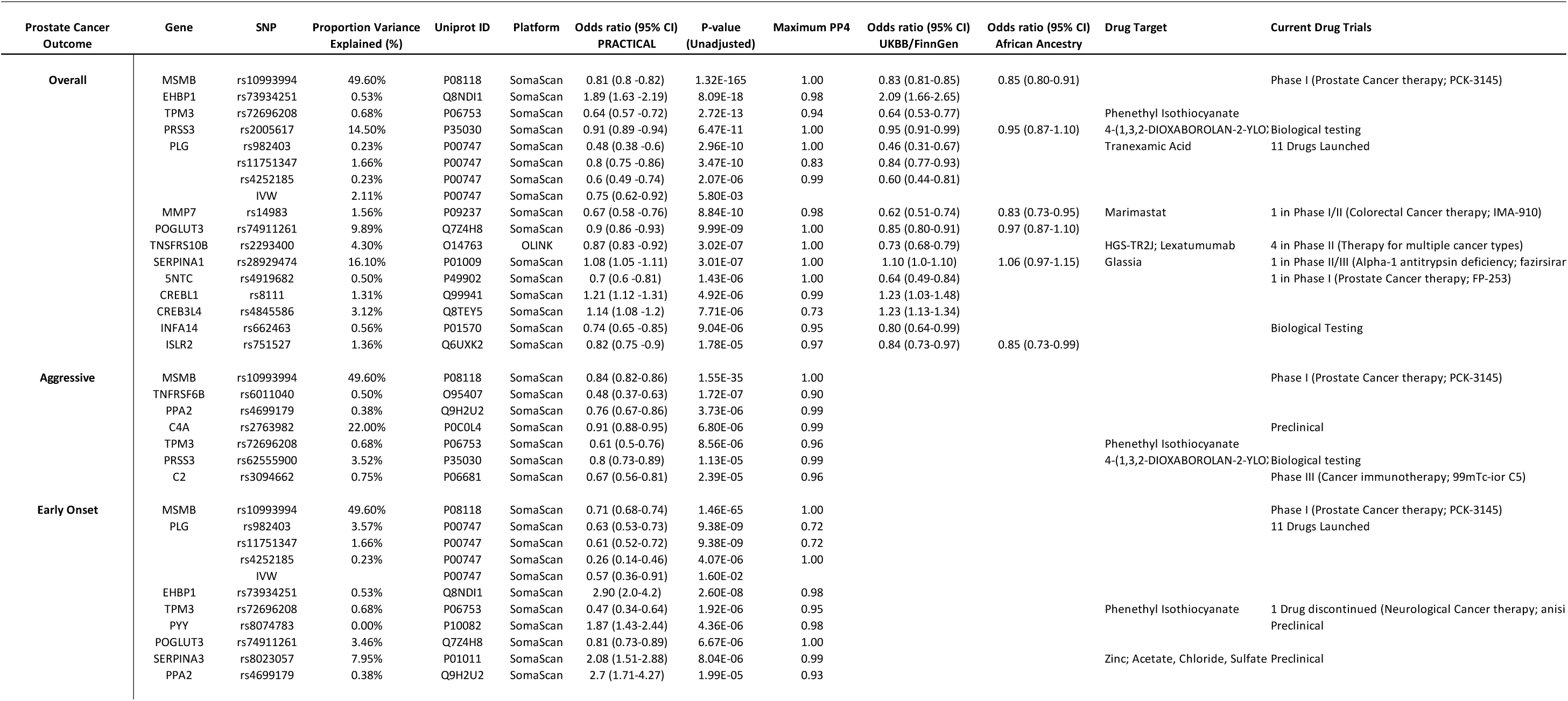
Mendelian randomization and colocalization results for protein-cancer associations that passed Bonferroni correction (0.05/n proteins analyzed) and colocalized at PP4 > 0.70. Results are shown per SNP-cancer association, except for those proteins for which there were multiple SNPs that passed both multiple testing correction and colocalization, and for which the summary estimate using the Inverse Variance Weighted (IVW) method is provided. Odds ratios are oriented per standard deviation increase in genetically predicted protein level. Maximum PP4 is reported as highest PP4 value from either single or conditional iterative colocalization method. Odds ratios are reported for associations using external UK Biobank/Finn Gen and for African ancestry population where data existed. Drug targets and drug trials are annotated if they existed.

The most statistically significant associations per standard deviation increase in protein level with evidence of colocalization were seen for MSMB (a protein that is specifically expressed in the prostate) with a lower risk of all prostate cancer endpoints [OR_Overall_= 0.81, 95% CI: 0.79-0.82, PP4: 100%; OR_Aggressive_ = 0.84, 95% CI: 0.82-0.86, PP4: 0.99; OR_Early Onset_ = 0.71, 95% CI: 0.68-0.74, PP4: 1.0, Table 1, Figure 2.]. TPM3 was the only other protein that had a colocalised association with risk of all outcomes [OR_Overall_ = 0.64, 95% CI: 0.57-0.73, PP4: 0.94; OR_Aggressive_= 0.61, 95% CI: 0.49-0.76, PP4: 0.96; OR_Early Onset_ = 0.47, 95% CI: 0.34-0.64, PP4: 0.95, Figure 2].

We also reported proteins with a colocalized association for one outcome and little evidence for an association with others after correction for multiple testing, such as IFNA14, ISLR2, MMP7, and TNSFRS10B which were associated exclusively with overall prostate cancer [OR_IFNA14_= 0.74, 95% CI: 0.70-0.78; OR_ISLR2_= 0.82, 95% CI: 0.75-0.90; OR_MMP7_= 0.67, 95% CI: 0.58-0.76; OR_TNFRSf10B_= 0.87, 95% CI: 0.83-0.92, Figure 2]. Similarly, PYY and SERPINA3 associated with an increased risk of early onset prostate cancer only [OR_PYY_ = 1.87, 95% CI: 1.43-2.44; OR_SERPINA3_ = 2.08, 95% CI: 1.51-2.88, Figure 2] while C2 associated with aggressive prostate cancer only [OR_C2_ = 0.67, 95% CI: 0.56-0.81, Figure 2].

We additionally identified proteins with evidence for a directionally concordant colocalized association with some but not all prostate cancer outcomes, including TNFRSF6B that had an inverse association with all outcomes but only showed evidence in favor of colocalization for aggressive disease [OR_Overall_ = 0.53, 95% CI: 0.46-0.61, PP4: 0.00; OR_Aggressive_ = 0.48, 95% CI: 0.0.37-0.63, PP4: 0.90; OR_Early Onset_ = 0.43, 95% CI: 0.29-0.63, PP4: 0.14, Figure 2,

Supplementary Table 1]. Likewise PPA2 was associated with an increased risk of both aggressive and early onset disease but lacked support from colocalization analyses for prostate cancer risk overall [OR_Overall_ = 1.84, 95% CI: 1.52-2.22, PP4: 0.01; OR_Aggressive_ = 2.13, 95% CI: 1.54-2.93, PP4: 0.99; OR_Early Onset_ = 2.70, 95% CI: 1.71-4.27, PP4: 0.93, Figure 2, Supplementary Table 1].

### Replication of robust proteins in European and African ancestry populations

We replicated the association for all 14 proteins that were robustly associated with overall prostate cancer (5NTC, CREBL1, CREB3L4, EHBP1, INFA14, ISLR2, MMP7, MSMB, PRSS3, PLG, POGLUT3, SERPINA1, TNSFRF10B, TPM3) using an independent meta-analysis of European ancestry participants in the UK Biobank and FinnGen cohorts (Table 1). Among these, the most statistically significant association was for MSMB [OR_Overall_ = 0.83, 95% CI: 0.81 to 0.85, Table 1] and the largest effect size was for PLG [OR_Overall_ = 0.46, 95% CI: 0.07 to 0.84, Table 1]. We additionally identified African ancestry-specific *cis-*pQTL for six of the 14 proteins (ISLR2, MMP7, MSMB, POGLUT3, PRSS3, SERPINA1; Table 1). Of these, three proteins associations with risk of prostate cancer overall were replicated in men of African ancestry: MSMB [OR_Overall_ = 0.85, 95% CI: 0.80 to 0.91], MMP7 [OR_Overall_ = 0.83, 95% CI: 0.73 to 0.95], and ISLR2 [OR_Overall_ = 0.85, 95% CI: 0.73 to 0.99] (Figure 3, Table 1).

**Figure 3.**
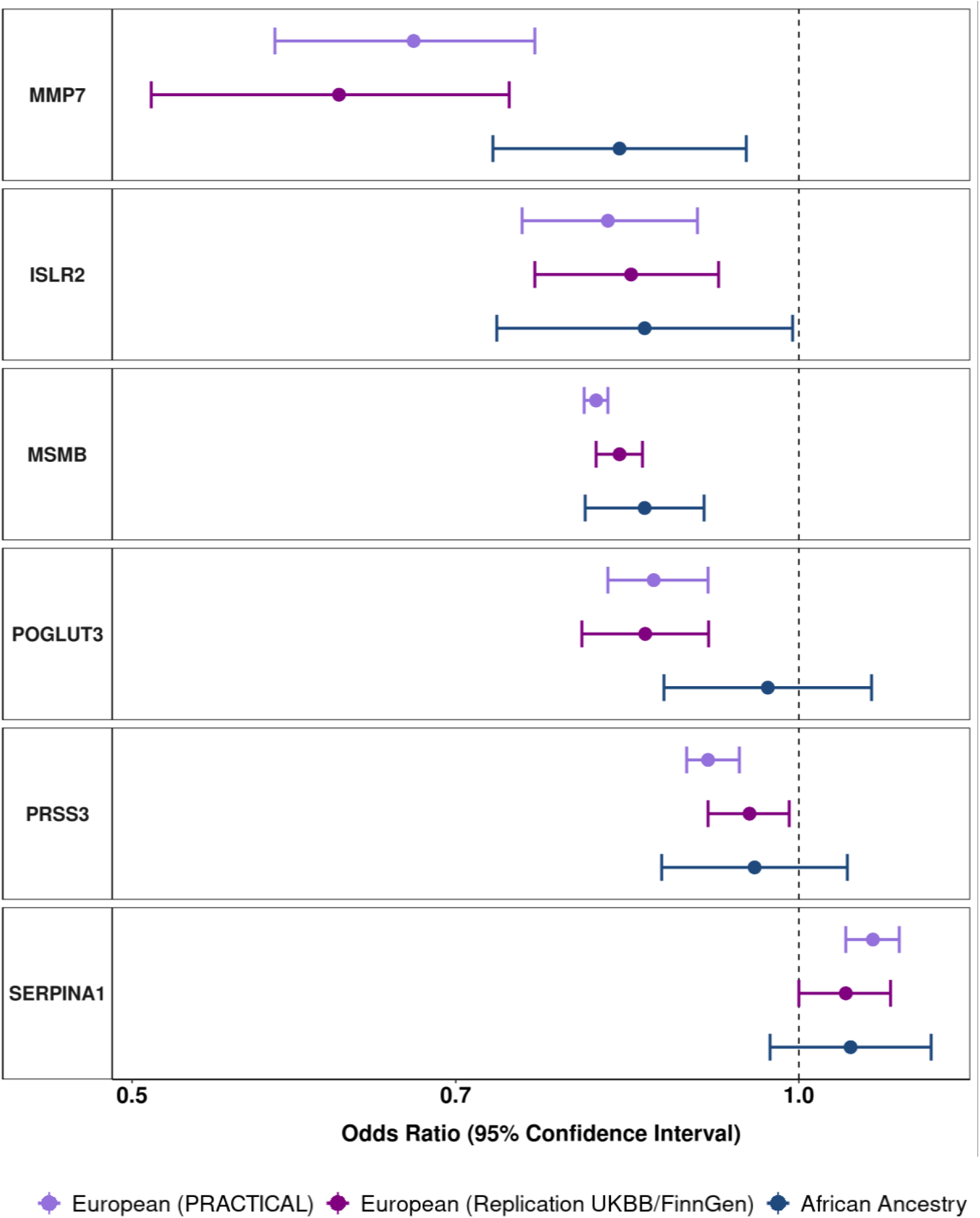
Odds ratios (95% confidence intervals) for genetically predicted protein levels and overall prostate cancer risk for proteins with p< Bonferroni threshold based on 0.05/number of proteins analyzed in main analyses, and with data available to perform replication in an African ancestry and European ancestry population. Odds ratio estimates are scaled per standard deviation increment in genetically predicted circulating protein concentrations.

### Drug target analysis

Out of the 2,002 unique proteins investigated, we identified 525 proteins that could be successfully mapped at the gene level to the target of a therapeutic intervention. Of these, ten (TPM3, PRSS3, PLG, MMP7, SERPINA1, SERPINA3, TNFRSF10B, C4A, HDGF, and LAYN) were associated with risk of at least one prostate cancer outcome after correction for investigating 525 drug target proteins and had evidence of colocalization (Supplementary Table 1). For example, PLG and C4A mapped to the clot dissolving class of fibrinolytics, TPM3 mapped to phenethyl isothiocyanate, and MMP7 mapped to matrix metalloproteinase inhibitor, marimastat.

### Spatial transcriptomic analysis

Of the 20 proteins that were associated with at least once prostate cancer outcome, we performed a targeted follow-up analysis for the two proteins with known high expression in the prostate tumor epithelium, MSMB and CREB3L4, using organ-wide spatial transcriptomic data on tissue obtained by radical prostatectomy from a patient with multifocal prostate cancer.^34^ In analyzing epithelial-rich spots, we observed marked differences of MSMB expression between benign cells, where MSMB was highly abundant, and Gleason grade group 4 (GG4) cells, where MSMB was very low or absent in a majority of cells (log MSMB_benign_ [median, interquartile range]: 2.73 [2.00-3.12] vs. log MSMB_GG4_: 0.48 [0.00-1.20], Figure 4.). A similarly, albeit more modestly, lower MSMB expression was observed in GG1 and GG2 cells (log MSMB_GG1_: 1.62 [0.97-1.88] and log MSMB_GG2_: 0.95 [0.69-1.32] compared to benign cells (Figure 4.). A lower expression of CREB3L4 was noted in GG2 and GG4 cells compared to benign cells (log CREB3L4_benign_: 0.60 [0.30-0.85] vs. log CREB3L4_GG2_: 0.31 [0.00-0.48] vs. log CREB3L4_GG4_: 0.48 [0.00-0.85], Supplementary Figure 2). Additional genome-wide random forest analyses identified MSMB expression as the most important gene in terms of distinguishing between benign and GG4 cells (Figure 5).

**Figure 4.**
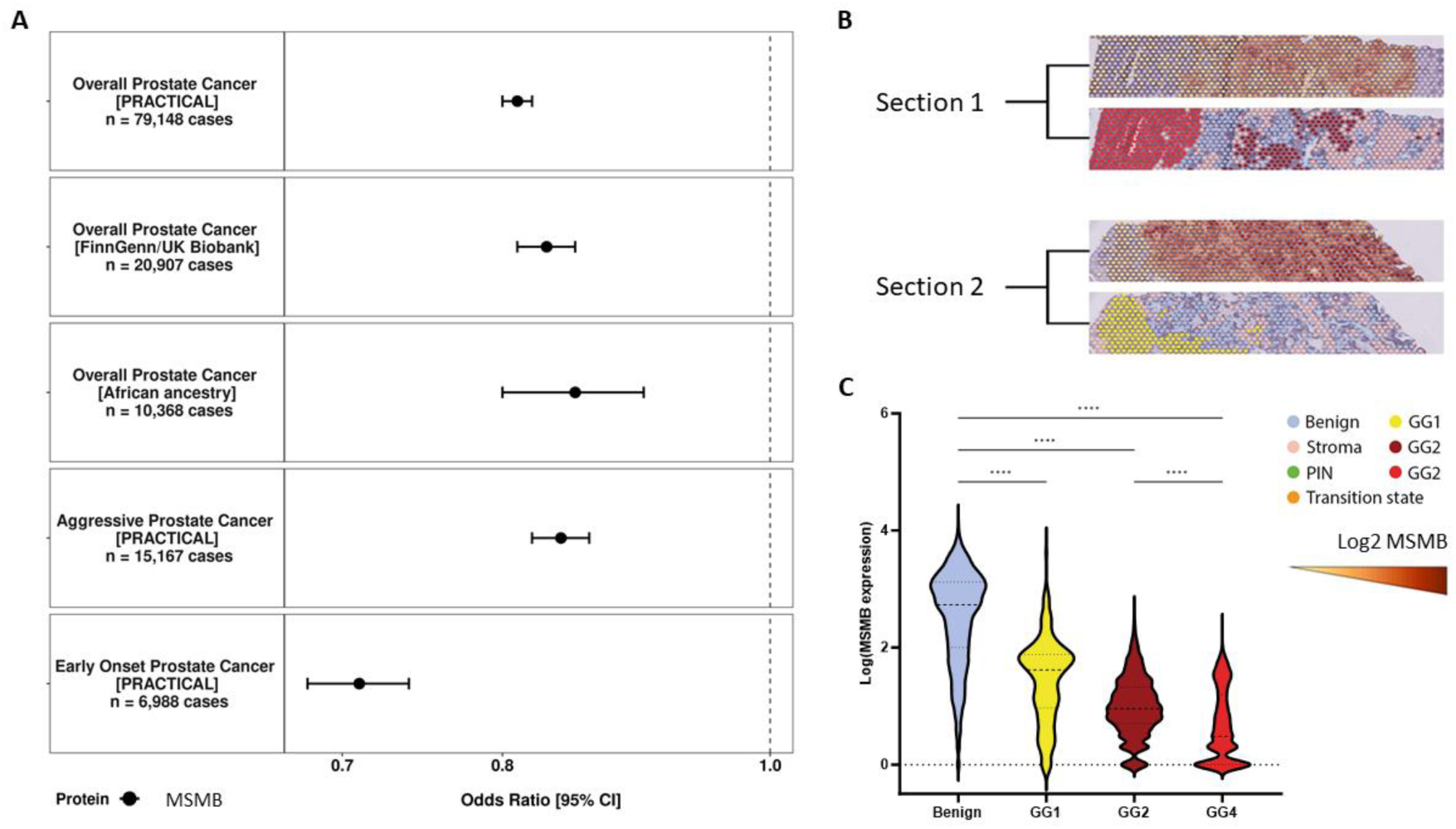
**A)** MSMB association with overall, early onset, and aggressive prostate cancer risk with replication in the FinnGen and UK Biobank populations and in an African ancestry population. Odds ratio (95% confidence interval) estimates are scaled per standard deviation increment in genetically predicted circulating MSMB concentrations **B)** Spatial visualization showing MSMB gene expression (top) and histology and tissue status (bottom) from organ-wide spatial transcriptomic data in two tumor sections (GG: Gleason grade group: GG1, Gleason score of 6 or lower; GG2, Gleason score of 3+4 = 7; GG4, Gleason score of 8). **C)** Violin plots representing gene expression in each spatial transcriptomics spot according to histological status. Statistical differences are indicated: **** p < 0.0001 (Kruskal–Wallis; post-test: Dunn’s test).

**Figure 5.**
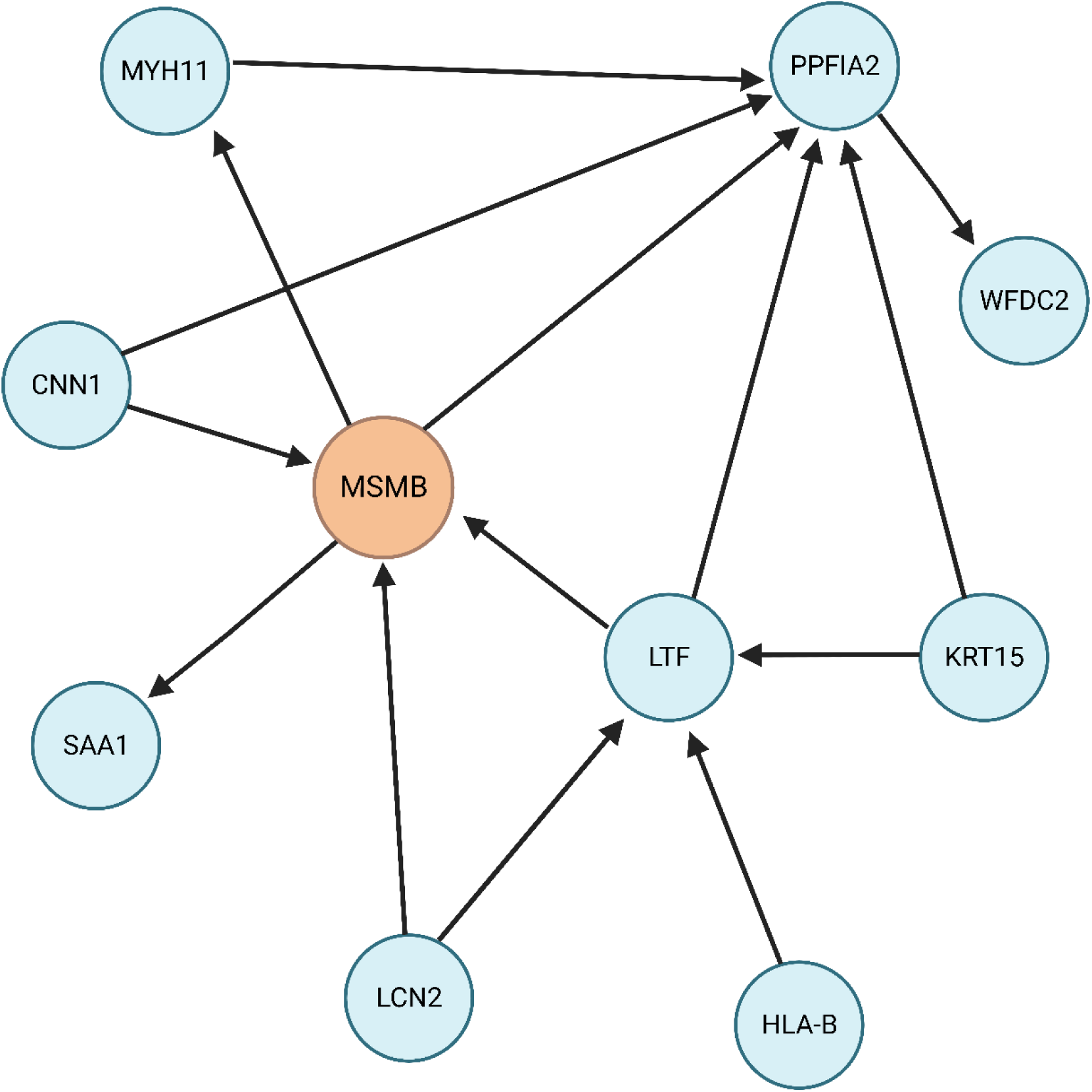
Gene network from iterative random forests of the difference in gene expression between benign and GG4 prostate histology (Gleason Score = 8). Arrows indicate direction of influence and shape of the network. MSMB is colored to demonstrate its central role in the network.

## Discussion

In this analysis, we conducted the largest study to date investigating the genetic associations of up to 2,002 unique proteins with the likely etiological risk of overall, early onset, and aggressive prostate cancer in up to 177,526 men using a MR and colocalization pipeline. In total, we found evidence supporting associations between 20 proteins and prostate cancer risk: 14 proteins for overall prostate cancer risk, seven for aggressive prostate cancer and eight for early onset prostate cancer. Among those 14 proteins that associated with prostate cancer risk overall, 14 were replicated in an external European ancestry population and three (out of six with available data) were replicated in an African ancestry population. A further half of the 20 proteins identified were also found to be the site of action for established drug targets with potential therapeutic implications. Finally, using spatial transcriptomics, we demonstrated a central role for the gene of our most robustly associated prostate cancer protein, MSMB, in distinguishing benign from undifferentiated, high-grade prostate cancer cells.

### MSMB

MSMB is a secretory protein and member of the human immunoglobulin family that is released largely by luminal epithelial cells in the prostate epithelium and has a documented role in overall prostate cancer risk for both observational and genetic epidemiology.^6,7,36^ In this study, we expand upon these previous findings by demonstrating that a 20% lower risk of overall prostate cancer is associated with higher levels of genetically predicted MSMB in two independent European ancestry cancer GWAS, and confirm for the first time its protective role in both aggressive and early onset disease etiology. We further successfully replicated this association with prostate cancer risk overall in an African-ancestry population, an ancestry group with an established higher risk of developing the disease. Subsequently, in reporting that MSMB gene expression is significantly depleted among high grade tumor when compared to expression on benign cells, we reiterate, through an independent line of evidence, that this gene may be particularly relevant to tumorigenesis and risk for aggressive disease.

Although the mechanism of action for MSMB in prostate cancer is not clear, MSMB has been shown to have a regulatory effect on cell growth, which may be lost during tumorigenesis while a MSMB-derived polypeptide was shown to induce prostate cell death.^36,37^ Additionally, in a rodent model and in vitro, higher MSMB activity was found to suppress prostate tumor growth while a knockout of MSMB promoter/enhancer regions was characterized by tumor progression and metastases.^38,39^ Given the integration of several compelling lines of evidence presented in this paper with existing literature, further research is warranted to understand the precise functional role of MSMB in prostate cancer tumorigenesis, identify environmental and lifestyle determinants, and explore potential clinical utility.

### Transcription Factors CREBL34 and CREBL1

We identified novel proteins associated with overall, early onset, and aggressive prostate cancer etiology. These include the endoplasmic reticulum (ER) originating transcription factors CREB3L4 and CREBL1 (also known as ATF6B), which we find are associated with an increased risk of prostate cancer overall and that are expressed in the prostate epithelium.^33^ These transcription factors form part of a transcriptional factor network that regulates the function of the endoplasmic reticulum (ER) and the activity of the unfolded protein response (UPR). There is an established role for the UPR and heat-shock proteins in maintaining AR stability and supporting AR-dependent tumorigenesis.^40,41^ CREB3L4 has been shown to directly interact with AR in LNCaP cells to increase cellular proliferation and is abundantly expressed in prostate tumor tissue.^42,43^ Furthermore, evidence suggests that disruptions in CREB3L4 contribute to ER stress downstream initiation of the unfolded protein response. ^43,44^ In a previous study of differential gene expression in prostate tissue, CREB3L4 was identified as a member of a co-expression gene cluster enriched for a previously described metabolic pathway (hsa05215) in prostate cancer that may regulate apoptosis and cell proliferation.^45^ Interestingly, we found that CREB3L4 expression is lower in GG2 and GG4 cells as opposed to benign cells.

Previous studies have linked AR activation with members of the ATF6 family in LNCaP and PC3 cells, however these experiments have mostly focused on ATF6A.^46,47^ For example, a recent *in vivo* study showed that prostate cancer cells with ATF6A overexpression resisted cellular death by ferroptosis.^46^ In parallel, a previous MR study reported a lowered risk of prostate cancer overall with genetically elevated circulating ATF6A levels from *trans-*pQTL.^48^ Given the promising role of its paralog, and the increased risk we report here, targeted follow up of CREBL1 may prove valuable in characterizing the broader role of ER stress proteins and AR-dependent tumorigenesis.

### EHBP1

We observed a more than two-fold increased risk of early-onset prostate cancer associated with higher EHBP1, an adaptor protein with a key role in vesicular trafficking and actin reorganization.^49^ Variants in the EHBP1 intron have previously been associated with aggressive prostate cancer in a genetic association study the protein and is more highly expressed in prostate tumor tissue and may have a role in determining the invasiveness of PTEN-positive prostate cancer cells according to GWAS and expression data and in a cellular study.^50–52^ While mechanisms that may link EHBP1 to prostate cancer risk are not yet fully described, it has role as an effector molecule for Rab8 family members that modulate polarized membrane transport via actin reorganization and may have a role in the mechanism of action for atorvastatin.^50^

### Other early onset and aggressive disease proteins

We found several proteins that were associated with early onset and aggressive disease, including PYY, PRSS3, PPA2, C2, C4A, and SERPINA3. For example, PYY is a metabolic hormone involved in appetite regulation, and while there has been some hypothesized relationship between obesity and aggressive prostate cancer risk in the past, recent findings suggest that obesity does not serve as a risk factor for disease itself, but may affect likelihood of diagnosis.^53,54^ Additionally, one study found that mesotrypsin, a protease encoded by the PRSS3 gene, was essential for prostate cancer metastasis in vitro and mouse models, however the role of this protein has not been widely studied in humans.^55^

We also note that proteins associated with early onset disease were generally greater in magnitude when compared to their associations with overall or aggressive disease. Two of these proteins, the complement proteins C2 and C4A, sit on chromosome six, which contains a particularly dense genetic region including the MHC complex and is consequently particularly difficult to interpret. However, given the importance of addressing early onset and aggressive disease, future studies are needed to further investigate and replicate the associations with early onset disease to uncover potential subtype specific mechanisms of disease onset and progression.

### Drug target proteins

We identified 10 proteins that were associated with both the risk of prostate cancer and that were the site of action for a known drug. These included TNFRSF10B, which is a receptor for the cytotoxic TRAIL ligand, and is essential for CASP8 and ER stress induced apoptosis.^56^ Further, TNFRSF10B expression is lower in higher grade prostate tumors and a recent study of PARP inhibitors in prostate cancer cell lines suggested that TNFRSF10B may provide a mechanism by which the cancer drug Olaparib induces apoptosis.^57^ The apparent protective association we observe with prostate cancer risk is in line with the results from multiple phase I/II trials of TNFRSF10B agonists that support their use for the treatment of multiple cancer endpoints, though not yet including prostate cancer. ^57–59^

We also identified an inverse association of PLG, a serine protease targeted by transexamic acids and several classes of thrombolytics, with prostate cancer risk overall and with early onset disease. Transexamic acids are primarily prescribed to control excessive bleeding while thrombolytics are primarily used to dissolve blot clots and act via plasmin and fibrin pathways.^60^ One molecular study found that PLG is generated by the cancer-mediated proteolysis of plasminogen which is released by human prostate carcinoma cells.^61^ PLG in turn has been shown in many lab studies to inhibit angiogenesis which when unregulated can lead to the rapid formation of tumors.^62,63^ Currently, several studies investigated combination therapy including plasminogen activation or inhibition for treatment of several cancer types, though not specifically for prostate cancer, in phase I/II trials.^64,65^

SERPINA1 maps to fazisiran, the treatment for alpha-1 antitrypsin deficiency and that is in phase II/III of drug trials. Previous findings have indicated that alpha-1 antitrypsin levels are often elevated in many carcinomas, including prostate.^66^ However, no agents targeting SERPINA1 have been investigated in cancer trials thus far. MMP7 belongs to a class of matrix metalloproteinases that participate in wound healing, bone growth, and matrix remodeling. There are multiple lines of evidence that this protein is involved in many cancers, and agents targeting metalloproteases, such as marimastat, are currently being investigated in clinical trials at various phases.^67^ In prostate cancer, marimastat showed some efficacy in early trials, however has not yet progressed further.^68^

While we highlight proteins that may share the same target site for established drug targets that may have implications for therapeutic use, the suitability of these to act as preventative or remedial agents requires careful considerations including the site specificity, potential downstream effects, routes of administration and effectively capturing the population at risk.^69^

### Strengths and weaknesses

This study offers several strengths including being the largest currently available GWAS of prostate cancer outcomes and the use of both aggressive and early onset endpoints with *cis-* pQTL covering up to 2,002 proteins. One previous MR study investigated the role of the circulating proteome in prostate cancer risk but did not stratify analyses by *cis* or *trans-* pQTL, and did not perform colocalization analyses, making it more challenging to infer causal relationships between individual proteins and cancer risk.^48^ Additionally, by integrating gene expression data measured using spatial transcriptomics, the current paper introduces a novel translational approach to highlight biological enablers of prostate cancer. To our knowledge, this study provides the first demonstration that MR using *cis* instruments of plasma protein levels can be used to identify a risk protein that has both specific expression in the cell of cancer origin and is related to tumor aggressiveness – important features to consider when identifying candidate targets for therapeutic prevention.

While we have analyzed a wide array of proteins, we have not investigated the entire human plasma proteome (n ∼ 20,000 protein-coding genes). As more protein GWAS data become available, it will become possible to use genetic methods to investigate more proteins. However, some blood proteins are unlikely to have a *cis-*pQTL due to the degree of evolutionary constraint for a protein’s cognate gene. Additional limitations include the more modest GWAS sample sizes for aggressive and early onset prostate cancer, which have lower power to discover novel protein associations. Finally, while we were able to perform additional analyses to replicate some, but not all, of our robust proteins in populations of African ancestry, we note as a limitation that GWAS sample sizes in this group are not yet sufficient to perform well-powered discovery analyses. Especially given the increased risk for prostate cancer among populations of African ancestry, it is essential that future studies identify risk proteins in more diverse populations and allow for the discovery of ancestry-specific markers of risk.

## Conclusion

This paper provides a catalogue of 20 proteins with evidence of etiological significance for prostate cancer. These proteins present an opportunity to direct further molecular and epidemiological investigations aimed at exploring the specific roles that the proteome plays in tumorigenesis and ultimately may inform future research into therapeutic prevention. In particular, converging evidence from population genetic and tumor sequencing analyses implicates MSMB as having an important protective role in prostate tumorigenesis, both in European and African-ancestry men, which is particularly marked for aggressive and early onset disease.

## Data Availability

Exposure data are publicly available through previously published and cited papers. Outcome data are available upon application and approval from the PRACTICAL Consortium.

## Disclaimer

Where authors are identified as personnel of the International Agency for Research on Cancer / World Health Organization, the authors alone are responsible for the views expressed in this article and they do not necessarily represent the decisions, policy, or views of the International Agency for Research on Cancer / World Health Organization. The findings and conclusions in this article are those of the authors and do not necessarily represent the official position of the National Institutes of Health. Department of Health and Social Care disclaimer: The views expressed are those of the author(s) and not necessarily those of the NHS, the NIHR or the Department of Health and Social Care. Anders Malarstig, Åsa Hedman, and Marios Dimitriou are employees of Pfizer Inc.

## Supplementary material. PRACTICAL Authors Information

### PIs from the PRACTICAL (http://practical.icr.ac.uk/), CRUK, BPC3, CAPS, PEGASUS consortia

Rosalind A. Eeles^1,2^, Christopher A. Haiman^3^, Zsofia Kote-Jarai^1^, Fredrick R. Schumacher^4,5^, Sara Benlloch^6,1^, Ali Amin Al Olama^6,7^, Kenneth R. Muir^8^, Sonja I. Berndt^9^, David V. Conti^3^, Fredrik Wiklund^10^, Stephen Chanock^9^, Ying Wang^11^, Catherine M. Tangen^12^, Jyotsna Batra^13,14,15^, Judith A. Clements^13,14,15^, APCB BioResource (Australian Prostate Cancer BioResource)^16,17^, Henrik Grönberg^10^, Nora Pashayan^18,19^, Johanna Schleutker^20,21^, Demetrius Albanes^9^, Stephanie J. Weinstein^9^, Alicja Wolk^22^, Catharine M. L. West^23^, Lorelei A. Mucci^24^, Géraldine Cancel-Tassin^25,26^, Stella Koutros^9^, Karina Dalsgaard Sørensen^27,28^, Eli Marie Grindedal^29^, David E. Neal^30,31,32^, Freddie C. Hamdy^33,34^, Jenny L. Donovan^35^, Ruth C. Travis^36^, Robert J. Hamilton^37,38^, Sue Ann Ingles^39^, Barry S. Rosenstein^40^, Yong-Jie Lu^41^, Graham G. Giles^42,43,44^, Robert J. MacInnis^42,43^, Adam S. Kibel^45^, Ana Vega^46,47,48^, Manolis Kogevinas^49,50,51,52^, Kathryn L. Penney^53^, Jong Y. Park^54^, Janet L. Stanford^55,56^, Cezary Cybulski^57^, Børge G. Nordestgaard^58,59^, Sune F. Nielsen^58,59^, Hermann Brenner^60,61,62^, Christiane Maier^63^, Jeri Kim^64^, Esther M. John^65^, Manuel R. Teixeira^66,67,68^, Susan L. Neuhausen^69^, Kim De Ruyck^70^, Azad Razack^71^, Lisa F. Newcomb^55,72^, Davor Lessel^73^, Radka Kaneva^74^, Nawaid Usmani^75,76^, Frank Claessens^77^, Paul A. Townsend^78,79^, Jose Esteban Castelao^80^, Monique J. Roobol^81^, Florence Menegaux^82^, Kay-Tee Khaw^83^, Lisa Cannon-Albright^84,85^, Hardev Pandha^79^, Stephen N. Thibodeau^86^, David J. Hunter^87^, Peter Kraft^88^, William J. Blot^89,90^, Elio Riboli^91^

^1^The Institute of Cancer Research, London, SM2 5NG, UK

^2^Royal Marsden NHS Foundation Trust, London, SW3 6JJ, UK

^3^Center for Genetic Epidemiology, Department of Preventive Medicine, Keck School of Medicine, University of Southern California/Norris Comprehensive Cancer Center, Los Angeles, CA 90015, USA

^4^Department of Population and Quantitative Health Sciences, Case Western Reserve University, Cleveland, OH 44106-7219, USA

^5^Seidman Cancer Center, University Hospitals, Cleveland, OH 44106, USA.

^6^Centre for Cancer Genetic Epidemiology, Department of Public Health and Primary Care, University of Cambridge, Strangeways Research Laboratory, Cambridge CB1 8RN, UK 7University of Cambridge, Department of Clinical Neurosciences, Stroke Research Group, R3, Box 83, Cambridge Biomedical Campus, Cambridge CB2 0QQ, UK

^8^Division of Population Health, Health Services Research and Primary Care, University of Manchester, Oxford Road, Manchester, M13 9PL, UK

^9^Division of Cancer Epidemiology and Genetics, National Cancer Institute, NIH, Bethesda, Maryland, 20892, USA

^10^Department of Medical Epidemiology and Biostatistics, Karolinska Institute, SE-171 77 Stockholm, Sweden

^11^Department of Population Science, American Cancer Society, 250 Williams Street, Atlanta, GA 30303, USA

^12^SWOG Statistical Center, Fred Hutchinson Cancer Research Center, Seattle, WA 98109, USA

^13^School of Biomedical Sciences, Faculty of Health, Queensland University of Technology, Brisbane, Queensland, Australia

^14^Centre for Genomics and Personalised Health, Queensland University of Technology, Brisbane, Queensland, Australia

^15^Translational Research Institute, QUT, Woolloongabba, Brisbane, Queensland, Australia 16Australian Prostate Cancer Research Centre-Qld, Queensland University of Technology, Brisbane; Prostate Cancer Research Program, Monash University, Melbourne; Dame Roma Mitchell Cancer Centre, University of Adelaide, Adelaide; Chris O’Brien Lifehouse and The Kinghorn Cancer Centre, Sydney, Australia

^17^Translational Research Institute, Brisbane, Queensland 4102, Australia

^18^Department of Applied Health Research, University College London, London, WC1E 7HB, UK

^19^Centre for Cancer Genetic Epidemiology, Department of Oncology, University of Cambridge, Strangeways Laboratory, Worts Causeway, Cambridge, CB1 8RN, UK 20Institute of Biomedicine, University of Turku, Finland

^21^Department of Medical Genetics, Genomics, Laboratory Division, Turku University Hospital, PO Box 52, 20521 Turku, Finland

^22^Institute of Environmental Medicine, Karolinska Institutet, 177 77 Stockholm, Sweden 23Division of Cancer Sciences, University of Manchester, Manchester Academic Health Science Centre, Radiotherapy Related Research, The Christie Hospital NHS Foundation Trust, Manchester, M13 9PL UK

^24^Department of Epidemiology, Harvard T. H. Chan School of Public Health, Boston, MA 02115, USA

^25^CeRePP, Tenon Hospital, F-75020 Paris, France.

^26^Sorbonne Universite, GRC n°5, AP-HP, Tenon Hospital, 4 rue de la Chine, F-75020 Paris, France

^27^Department of Molecular Medicine, Aarhus University Hospital, Palle Juul-Jensen Boulevard 99, 8200 Aarhus N, Denmark

^28^Department of Clinical Medicine, Aarhus University, DK-8200 Aarhus N

^29^Department of Medical Genetics, Oslo University Hospital, 0424 Oslo, Norway

^30^Nuffield Department of Surgical Sciences, University of Oxford, Room 6603, Level 6, John Radcliffe Hospital, Headley Way, Headington, Oxford, OX3 9DU, UK

^31^University of Cambridge, Department of Oncology, Box 279, Addenbrooke’s Hospital, Hills Road, Cambridge CB2 0QQ, UK

^32^Cancer Research UK, Cambridge Research Institute, Li Ka Shing Centre, Cambridge, CB2 0RE, UK

^33^Nuffield Department of Surgical Sciences, University of Oxford, Oxford, OX1 2JD, UK 34Faculty of Medical Science, University of Oxford, John Radcliffe Hospital, Oxford, UK 35Population Health Sciences, Bristol Medical School, University of Bristol, BS8 2PS, UK 36Cancer Epidemiology Unit, Nuffield Department of Population Health, University of Oxford, Oxford, OX3 7LF, UK

^37^Dept. of Surgical Oncology, Princess Margaret Cancer Centre, Toronto ON M5G 2M9, Canada

^38^Dept. of Surgery (Urology), University of Toronto, Canada

^39^Department of Preventive Medicine, Keck School of Medicine, University of Southern California/Norris Comprehensive Cancer Center, Los Angeles, CA 90015, USA 40Department of Radiation Oncology and Department of Genetics and Genomic Sciences, Box 1236, Icahn School of Medicine at Mount Sinai, One Gustave L. Levy Place, New York, NY 10029, USA

^41^Centre for Cancer Biomarker and Biotherapeutics, Barts Cancer Institute, Queen Mary University of London, John Vane Science Centre, Charterhouse Square, London, EC1M 6BQ, UK

^42^Cancer Epidemiology Division, Cancer Council Victoria, 200 Victoria Parade, East Melbourne, VIC, 3002, Australia

^43^Centre for Epidemiology and Biostatistics, Melbourne School of Population and Global Health, The University of Melbourne, Grattan Street, Parkville, VIC 3010, Australia 44Precision Medicine, School of Clinical Sciences at Monash Health, Monash University, Clayton, Victoria 3168, Australia

^45^Division of Urologic Surgery, Brigham and Womens Hospital, 75 Francis Street, Boston, MA 02115, USA

^46^Fundación Pública Galega Medicina Xenómica, Santiago de Compostela, 15706, Spain. 47Instituto de Investigación Sanitaria de Santiago de Compostela, Santiago de Compostela, 15706, Spain.

^48^Centro de Investigación en Red de Enfermedades Raras (CIBERER), Spain

^49^ISGlobal, Barcelona, Spain

^50^IMIM (Hospital del Mar Medical Research Institute), Barcelona, Spain

^51^Universitat Pompeu Fabra (UPF), Barcelona, Spain

^52^CIBER Epidemiología y Salud Pública (CIBERESP), 28029 Madrid, Spain

^53^Channing Division of Network Medicine, Department of Medicine, Brigham and Women’s Hospital/Harvard Medical School, Boston, MA 02115, USA

^54^Department of Cancer Epidemiology, Moffitt Cancer Center, 12902 Magnolia Drive, Tampa, FL 33612, USA

^55^Division of Public Health Sciences, Fred Hutchinson Cancer Research Center, Seattle, Washington, 98109-1024, USA

^56^Department of Epidemiology, School of Public Health, University of Washington, Seattle, Washington 98195, USA

^57^International Hereditary Cancer Center, Department of Genetics and Pathology, Pomeranian Medical University, 70-115 Szczecin, Poland

^58^Faculty of Health and Medical Sciences, University of Copenhagen, 2200 Copenhagen, Denmark

^59^Department of Clinical Biochemistry, Herlev and Gentofte Hospital, Copenhagen University Hospital, Herlev, 2200 Copenhagen, Denmark

^60^Division of Clinical Epidemiology and Aging Research, German Cancer Research Center (DKFZ), D-69120, Heidelberg, Germany

^61^German Cancer Consortium (DKTK), German Cancer Research Center (DKFZ), D-69120 Heidelberg, Germany

^62^Division of Preventive Oncology, German Cancer Research Center (DKFZ) and National Center for Tumor Diseases (NCT), Im Neuenheimer Feld 460, 69120 Heidelberg, Germany 63Humangenetik Tuebingen, Paul-Ehrlich-Str 23, D-72076 Tuebingen, Germany

^64^The University of Texas M. D. Anderson Cancer Center, Department of Genitourinary Medical Oncology, 1515 Holcombe Blvd., Houston, TX 77030, USA

^65^Departments of Epidemiology & Population Health and of Medicine, Division of Oncology, Stanford Cancer Institute, Stanford University School of Medicine, Stanford, CA 94304 USA 66Department of Laboratory Genetics, Portuguese Oncology Institute of Porto (IPO Porto) / Porto Comprehensive Cancer Center, Porto, Portugal

^67^Cancer Genetics Group, IPO Porto Research Center (CI-IPOP) / RISE@CI-IPOP (Health Research Network), Portuguese Oncology Institute of Porto (IPO Porto) / Porto Comprehensive Cancer Center, Porto, Portugal

^68^School of Medicine and Biomedical Sciences (ICBAS), University of Porto, Porto, Portugal 69Department of Population Sciences, Beckman Research Institute of the City of Hope, 1500 East Duarte Road, Duarte, CA 91010

^70^Ghent University, Faculty of Medicine and Health Sciences, Basic Medical Sciences, Proeftuinstraat 86, B-9000 Gent

^71^Department of Surgery, Faculty of Medicine, University of Malaya, 50603 Kuala Lumpur, Malaysia

^72^Department of Urology, University of Washington, 1959 NE Pacific Street, Box 356510, Seattle, WA 98195, USA

^73^Institute of Human Genetics, University Medical Center Hamburg-Eppendorf, D-20246 Hamburg, Germany

^74^Molecular Medicine Center, Department of Medical Chemistry and Biochemistry, Medical University of Sofia, Sofia, 2 Zdrave Str., 1431 Sofia, Bulgaria

^75^Department of Oncology, Cross Cancer Institute, University of Alberta, 11560 University Avenue, Edmonton, Alberta, Canada T6G 1Z2

^76^Division of Radiation Oncology, Cross Cancer Institute, 11560 University Avenue, Edmonton, Alberta, Canada T6G 1Z2

^77^Molecular Endocrinology Laboratory, Department of Cellular and Molecular Medicine, KU Leuven, BE-3000, Belgium

^78^Division of Cancer Sciences, Manchester Cancer Research Centre, Faculty of Biology, Medicine and Health, Manchester Academic Health Science Centre, NIHR Manchester Biomedical Research Centre, Health Innovation Manchester, Univeristy of Manchester, M13 9WL

^79^The University of Surrey, Guildford, Surrey, GU2 7XH, UK

^80^Genetic Oncology Unit, CHUVI Hospital, Complexo Hospitalario Universitario de Vigo, Instituto de Investigación Biomédica Galicia Sur (IISGS), 36204, Vigo (Pontevedra), Spain 81Department of Urology, Erasmus University Medical Center, Cancer Institute, 3015 GD Rotterdam, The Netherlands

^82^“Exposome and Heredity”, CESP (UMR 1018), Faculté de Médecine, Université Paris-Saclay, Inserm, Gustave Roussy, Villejuif

^83^Clinical Gerontology Unit, University of Cambridge, Cambridge, CB2 2QQ, UK

^84^Division of Epidemiology, Department of Internal Medicine, University of Utah School of Medicine, Salt Lake City, Utah 84132, USA

^85^George E. Wahlen Department of Veterans Affairs Medical Center, Salt Lake City, Utah 84148, USA

^86^Department of Laboratory Medicine and Pathology, Mayo Clinic, Rochester, MN 55905, USA

^87^Nuffield Department of Population Health, University of Oxford, United Kingdom 88Program in Genetic Epidemiology and Statistical Genetics, Department of Epidemiology, Harvard School of Public Health, Boston, MA, USA

^89^Division of Epidemiology, Department of Medicine, Vanderbilt University Medical Center, 2525 West End Avenue, Suite 800, Nashville, TN 37232 USA.

^90^International Epidemiology Institute, Rockville, MD 20850, USA

^91^Department of Epidemiology and Biostatistics, School of Public Health, Imperial College London, SW7 2AZ, UK

### CRUK and PRACTICAL consortium

This work was supported by the Canadian Institutes of Health Research, European Commission’s Seventh Framework Programme grant agreement n° 223175 (HEALTH-F2-2009-223175), Cancer Research UK Grants C5047/A7357, C1287/A10118, C1287/A16563, C5047/A3354, C5047/A10692, C16913/A6135, and The National Institute of Health (NIH) Cancer Post-Cancer GWAS initiative grant: No. 1 U19 CA 148537-01 (the GAME-ON initiative).

We would also like to thank the following for funding support: The Institute of Cancer Research and The Everyman Campaign, The Prostate Cancer Research Foundation, Prostate Research Campaign UK (now PCUK), The Orchid Cancer Appeal, Rosetrees Trust, The National Cancer Research Network UK, The National Cancer Research Institute (NCRI) UK, and Prostate Cancer Canada. We are grateful for support of NIHR funding to the NIHR Biomedical Research Centre at The Institute of Cancer Research, The Royal Marsden NHS Foundation Trust, and Manchester NIHR Biomedical Research Centre. The Prostate Cancer Program of Cancer Council Victoria also acknowledge grant support from The National Health and Medical Research Council, Australia (126402, 209057, 251533, 396414, 450104, 504700, 504702, 504715, 623204, 940394, 614296,), VicHealth, Cancer Council Victoria, The Prostate Cancer Foundation of Australia, The Whitten Foundation, PricewaterhouseCoopers, and Tattersall’s. EAO, DMK, and EMK acknowledge the Intramural Program of the National Human Genome Research Institute for their support.

Genotyping of the OncoArray was funded by the US National Institutes of Health (NIH) [U19 CA 148537 for ELucidating Loci Involved in Prostate cancer SuscEptibility (ELLIPSE) project and X01HG007492 to the Center for Inherited Disease Research (CIDR) under contract number HHSN268201200008I]. Additional analytic support was provided by NIH NCI U01 CA188392 (PI: Schumacher).

Research reported in this publication also received support from the National Cancer Institute of the National Institutes of Health under Award Numbers U10 CA37429 (CD Blanke), and UM1 CA182883 (CM Tangen/IM Thompson). The content is solely the responsibility of the authors and does not necessarily represent the official views of the National Institutes of Health.

Funding for the iCOGS infrastructure came from: the European Community’s Seventh Framework Programme under grant agreement n° 223175 (HEALTH-F2-2009-223175) (COGS), Cancer Research UK (C1287/A10118, C1287/A 10710, C12292/A11174, C1281/A12014, C5047/A8384, C5047/A15007, C5047/A10692, C8197/A16565), the National

Institutes of Health (CA128978) and Post-Cancer GWAS initiative (1U19 CA148537, 1U19 CA148065 and 1U19 CA148112 - the GAME-ON initiative), the Department of Defence (W81XWH-10-1-0341), the Canadian Institutes of Health Research (CIHR) for the CIHR Team in Familial Risks of Breast Cancer, Komen Foundation for the Cure, the Breast Cancer Research Foundation, and the Ovarian Cancer Research Fund

### BPC3

The BPC3 was supported by the U.S. National Institutes of Health, National Cancer Institute (cooperative agreements U01-CA98233 to D.J.H., U01-CA98710 to S.M.G., U01-CA98216 to E.R., and U01-CA98758 to B.E.H., and Intramural Research Program of NIH/National Cancer Institute, Division of Cancer Epidemiology and Genetics).

### CAPS

CAPS GWAS study was supported by the Cancer Risk Prediction Center (CRisP; www.crispcenter.org), a Linneus Centre (Contract ID 70867902) financed by the Swedish Research Council, (grant no K2010-70X-20430-04-3), the Swedish Cancer Foundation (grant no 09-0677), the Hedlund Foundation, the Soederberg Foundation, the Enqvist Foundation, ALF funds from the Stockholm County Council. Stiftelsen Johanna Hagstrand och Sigfrid Linner’s Minne, Karlsson’s Fund for urological and surgical research.

### PEGASUS

PEGASUS was supported by the Intramural Research Program, Division of Cancer Epidemiology and Genetics, National Cancer Institute, National Institutes of Health.

---

**Supplementary Figure 1.**
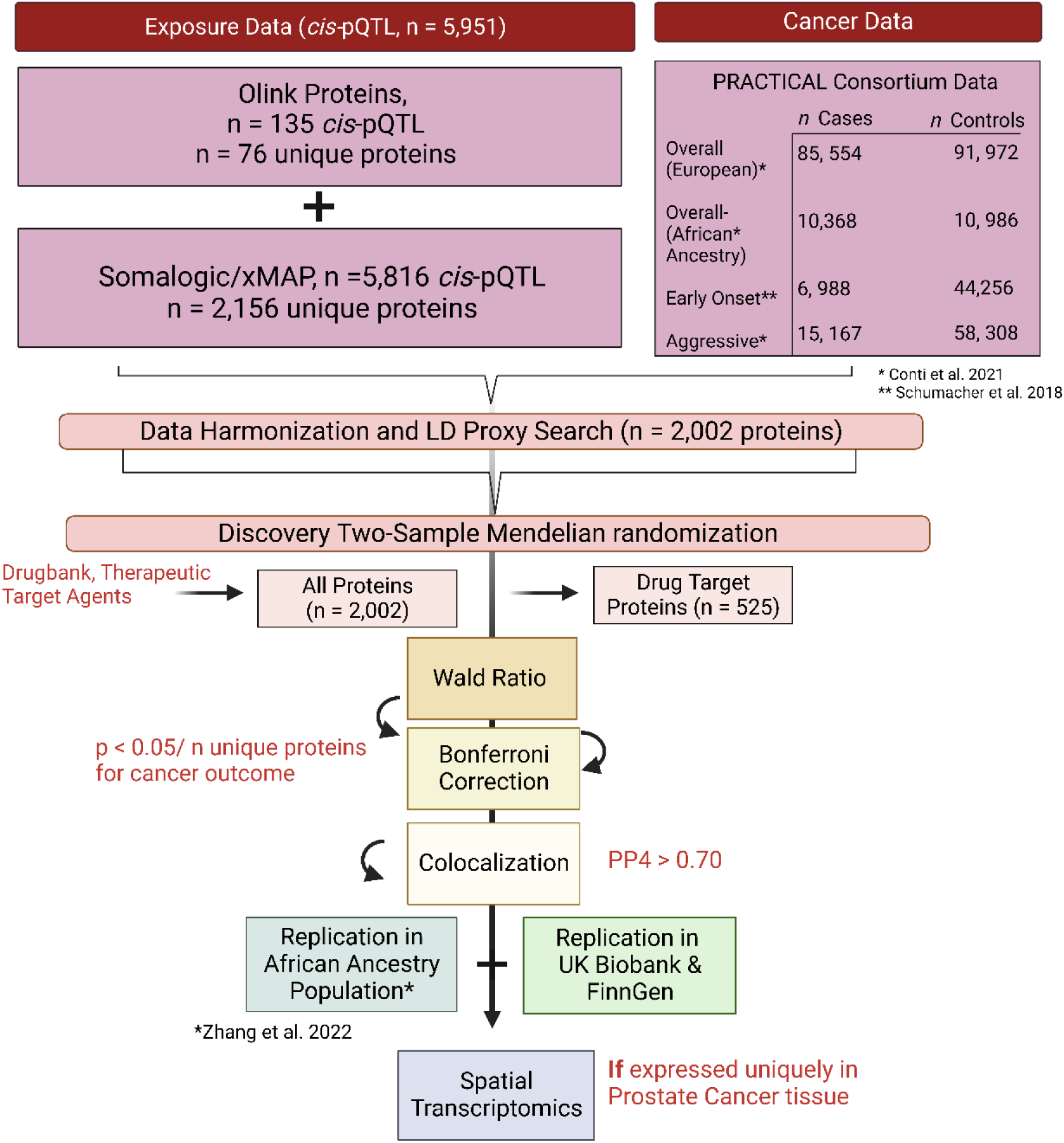
Flow-chart showing the overall study design. PP4 = posterior probability of a shared causal locus. LD = Linkage disequilibrium.

**Supplementary Figure 2.**
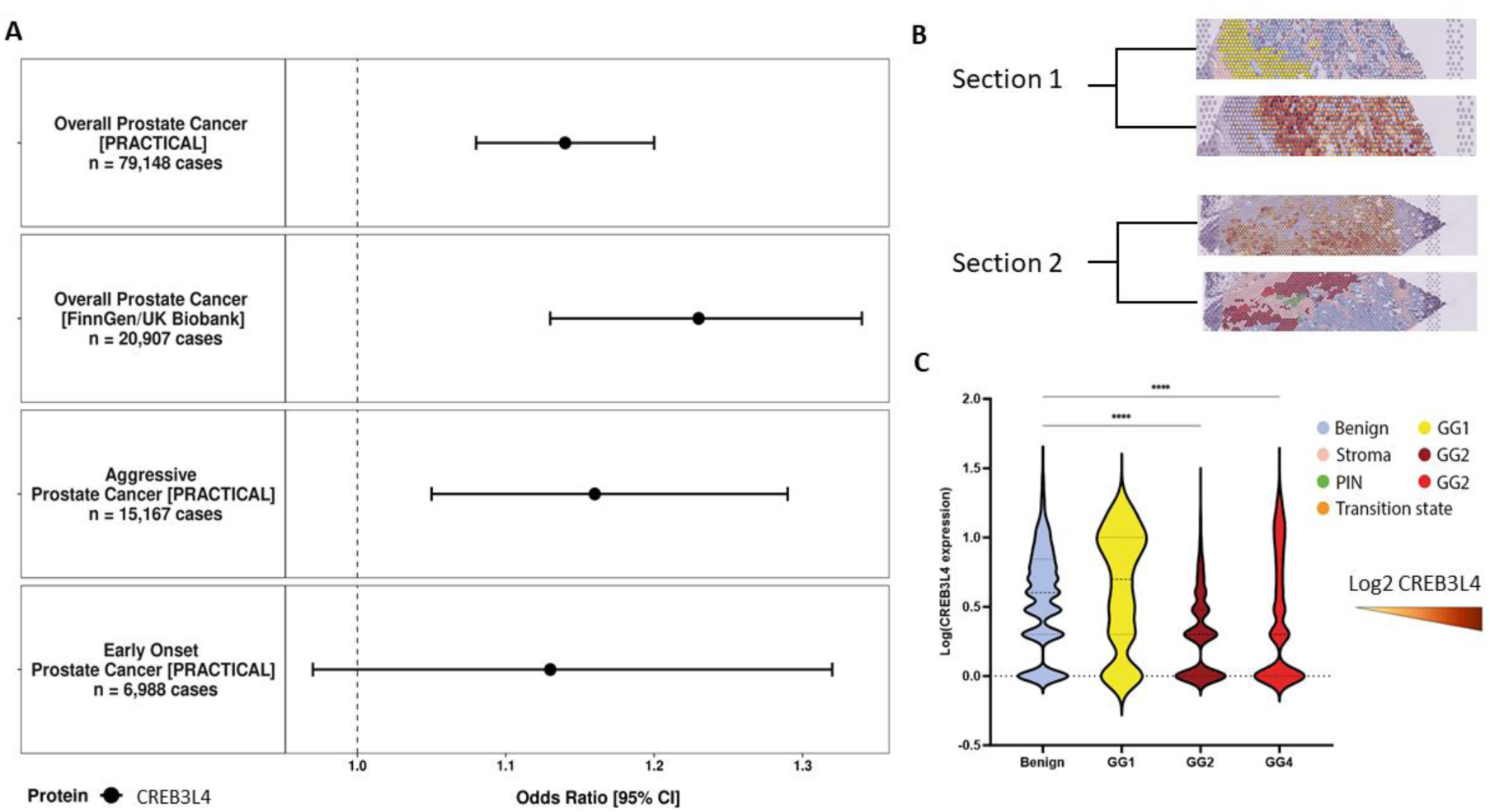
**A)** CREB3L4 association with overall, early onset, and aggressive prostate cancer risk with replication in the FinnGen and UK Biobank populations. Odds ratio (95% confidence interval) estimates are scaled per standard deviation increment in genetically predicted circulating CREB3L4 concentrations **B)** Spatial visualization showing CREB3L4 gene expression (top) and histology and tissue status (bottom) from organ-wide spatial transcriptomic data in two tumor sections (GG: Gleason grade group: GG1, Gleason score of 6 or lower; GG2, Gleason score of 3+4 = 7; GG4, Gleason score of 8). **C)** Violin plots representing gene expression in each spatial transcriptomics spot according to histological status. Statistical differences are indicated: **** p < 0.0001 (Kruskal–Wallis; post-test: Dunn’s test).

**Supplementary Table 1**.

Full protein data characteristic data and Mendelian randomization and colocalization results for all proteins and cancer outcomes, where analyzed. Odds ratios (95% confidence intervals) are given per standard deviation increase in genetically predicted protein level, and maximum colocalization indicates the highest PP4 percentage from either single or conditional iterative colocalization methods.

## References

1. Siegel, R. L., Miller, K. D. & Jemal, A. Cancer statistics, 2018. CA: A Cancer Journal for Clinicians 68, 7–30 (2018).

2. Salinas, C. A., Tsodikov, A., Ishak-Howard, M. & Cooney, K. A. Prostate Cancer in Young Men: An Important Clinical Entity. Nat Rev Urol 11, 317–323 (2014).

3. Shih, H.-J., Fang, S.-C., An, L. & Shao, Y.-H. J. Early-onset prostate cancer is associated with increased risks of disease progression and cancer-specific mortality. Prostate 81, 118–126 (2021).

4. Bergengren, O. et al. 2022 Update on Prostate Cancer Epidemiology and Risk Factors—A Systematic Review. European Urology 84, 191–206 (2023).

5. Watts, E. L. et al. Circulating insulin-like growth factors and risks of overall, aggressive and early-onset prostate cancer: a collaborative analysis of 20 prospective studies and Mendelian randomization analysis. International Journal of Epidemiology 52, 71–86 (2023).

6. Smith Byrne, K., et al. The role of plasma microseminoprotein-beta in prostate cancer: an observational nested case-control and Mendelian randomization study in the European prospective investigation into cancer and nutrition. Ann Oncol 30, 983–989 (2019).

7. Haiman, C. A. et al. Levels of Beta-Microseminoprotein in Blood and Risk of Prostate Cancer in Multiple Populations. J Natl Cancer Inst 105, 237–243 (2013).

8. Swerdlow, D. I. et al. Selecting instruments for Mendelian randomization in the wake of genome-wide association studies. Int J Epidemiol 45, 1600–1616 (2016).

9. Burgess, S., Foley, C. N. & Zuber, V. Inferring Causal Relationships Between Risk Factors and Outcomes from Genome-Wide Association Study Data. Annual Review of Genomics and Human Genetics 19, 303–327 (2018).

10. Giambartolomei, C. et al. Bayesian Test for Colocalisation between Pairs of Genetic Association Studies Using Summary Statistics. PLoS Genet 10, e1004383 (2014).

11. Lawlor, D. A., Tilling, K. & Davey Smith, G. Triangulation in aetiological epidemiology. Int. J. Epidemiol. dyw314 (2017) doi:10.1093/ije/dyw314.

12. Williams, C. G., Lee, H. J., Asatsuma, T., Vento-Tormo, R. & Haque, A. An introduction to spatial transcriptomics for biomedical research. Genome Medicine 14, 68 (2022).

13. Ferkingstad, E. et al. Large-scale integration of the plasma proteome with genetics and disease. Nat Genet 53, 1712–1721 (2021).

14. Zheng, J. et al. Phenome-wide Mendelian randomization mapping the influence of the plasma proteome on complex diseases. Nat Genet 52, 1122–1131 (2020).

15. Folkersen, L. et al. Genomic and drug target evaluation of 90 cardiovascular proteins in 30,931 individuals. Nat Metab 2, 1135–1148 (2020).

16. Sun, X. et al. Genetically predicted levels of circulating cytokines and prostate cancer risk: A Mendelian randomization study. Int. J. Cancer 147, 2469–2478 (2020).

17. Hemani, G. et al. The MR-Base platform supports systematic causal inference across the human phenome. eLife 7, e34408 (2018).

18. Fauman, E. B. & Hyde, C. An optimal variant to gene distance window derived from an empirical definition of cis and trans protein QTLs. BMC Bioinformatics 23, 169 (2022).

19. 19. PRACTICAL. http://practical.icr.ac.uk/.

20. Schumacher, F. R. et al. Association analyses of more than 140,000 men identify 63 new prostate cancer susceptibility loci. Nat Genet 50, 928–936 (2018).

21. Conti, D. V. et al. Trans-ancestry genome-wide association meta-analysis of prostate cancer identifies new susceptibility loci and informs genetic risk prediction. Nat Genet 53, 65–75 (2021).

22. Burgess, S., Foley, C. N., Allara, E., Staley, J. R. & Howson, J. M. M. A robust and efficient method for Mendelian randomization with hundreds of genetic variants. Nat Commun 11, 376 (2020).

23. Wallace, C. A more accurate method for colocalisation analysis allowing for multiple causal variants. PLOS Genetics 17, e1009440 (2021).

24. Wallace, C. Eliciting priors and relaxing the single causal variant assumption in colocalisation analyses. PLoS Genet 16, e1008720 (2020).

25. Kurki, M. I. et al. FinnGen provides genetic insights from a well-phenotyped isolated population. Nature 613, 508–518 (2023).

26. Hewitt, J., Walters, M., Padmanabhan, S. & Dawson, J. Cohort profile of the UK Biobank: diagnosis and characteristics of cerebrovascular disease. BMJ Open 6, e009161 (2016).

27. Zhang, J. et al. Plasma proteome analyses in individuals of European and African ancestry identify cis-pQTLs and models for proteome-wide association studies. Nat Genet 54, 593–602 (2022).

28. Wishart, D. S. et al. DrugBank 5.0: a major update to the DrugBank database for 2018. Nucleic Acids Res 46, D1074–D1082 (2018).

29. Sheils, T. K. et al. TCRD and Pharos 2021: mining the human proteome for disease biology. Nucleic Acids Res 49, D1334–D1346 (2021).

30. Zhou, Y. et al. Therapeutic target database update 2022: facilitating drug discovery with enriched comparative data of targeted agents. Nucleic Acids Research 50, D1398– D1407 (2022).

31. Erickson, A. et al. A Systematic Review of Prostate Cancer Heterogeneity: Understanding the Clonal Ancestry of Multifocal Disease. European Urology Oncology 4, 358–369 (2021).

32. Hunter, M. V., Moncada, R., Weiss, J. M., Yanai, I. & White, R. M. Spatially resolved transcriptomics reveals the architecture of the tumor-microenvironment interface. Nat Commun 12, 6278 (2021).

33. THE TABULA SAPIENS CONSORTIUM. The Tabula Sapiens: A multiple-organ, single-cell transcriptomic atlas of humans. Science 376, eabl4896 (2022).

34. Erickson, A. et al. Spatially resolved clonal copy number alterations in benign and malignant tissue. Nature 608, 360–367 (2022).

35. Basu, S., Kumbier, K., Brown, J. B. & Yu, B. Iterative random forests to discover predictive and stable high-order interactions. Proceedings of the National Academy of Sciences 115, 1943–1948 (2018).

36. Whitaker, H. C., Warren, A. Y., Eeles, R., Kote-Jarai, Z. & Neal, D. E. The potential value of microseminoprotein-beta as a prostate cancer biomarker and therapeutic target. Prostate 70, 333–340 (2010).

37. Annabi, B. et al. Contribution of the 37-kDa laminin receptor precursor in the anti-metastatic PSP94-derived peptide PCK3145 cell surface binding. Biochem Biophys Res Commun 346, 358–366 (2006).

38. Annabi, B. et al. A PSP94-derived peptide PCK3145 inhibits MMP-9 secretion and triggers CD44 cell surface shedding: implication in tumor metastasis. Clin Exp Metastasis 22, 429–439 (2005).

39. Lamy, S. et al. A prostate secretory protein94-derived synthetic peptide PCK3145 inhibits VEGF signalling in endothelial cells: implication in tumor angiogenesis. Int J Cancer 118, 2350–2358 (2006).

40. Jin, Y. & Saatcioglu, F. Targeting the Unfolded Protein Response in Hormone-Regulated Cancers. Trends in Cancer 6, 160–171 (2020).

41. Centenera, M. M., Fitzpatrick, A. K., Tilley, W. D. & Butler, L. M. Hsp90: Still a viable target in prostate cancer. Biochimica et Biophysica Acta (BBA) - Reviews on Cancer 1835, 211–218 (2013).

42. Labrie, C. et al. Androgen-regulated transcription factor AIbZIP in prostate cancer. The Journal of Steroid Biochemistry and Molecular Biology 108, 237–244 (2008).

43. Kim, T.-H., Park, J.-M., Kim, M.-Y. & Ahn, Y.-H. The role of CREB3L4 in the proliferation of prostate cancer cells. Sci Rep 7, 45300 (2017).

44. Pu, Q. et al. The Novel Transcription Factor CREB3L4 Contributes to the Progression of Human Breast Carcinoma. J Mammary Gland Biol Neoplasia 25, 37–50 (2020).

45. Pidò, S., Ceddia, G. & Masseroli, M. Computational analysis of fused co-expression networks for the identification of candidate cancer gene biomarkers. NPJ Syst Biol Appl 7, 17 (2021).

46. Zhao, R. et al. ATF6α promotes prostate cancer progression by enhancing PLA2G4A-mediated arachidonic acid metabolism and protecting tumor cells against ferroptosis. Prostate 82, 617–629 (2022).

47. Zhou, H. et al. The functional implication of ATF6α in castration-resistant prostate cancer cells. The FASEB Journal 37, e22758 (2023).

48. Wu, L. et al. Analysis of over 140,000 European descendants identifies genetically-predicted blood protein biomarkers associated with prostate cancer risk. Cancer Res 79, 4592–4598 (2019).

49. Rai, A., Bleimling, N., Vetter, I. R. & Goody, R. S. The mechanism of activation of the actin binding protein EHBP1 by Rab8 family members. Nat Commun 11, 4187 (2020).

50. Ghalali, A., Wiklund, F., Zheng, H., Stenius, U. & Högberg, J. Atorvastatin prevents ATP-driven invasiveness via P2X7 and EHBP1 signaling in PTEN-expressing prostate cancer cells. Carcinogenesis 35, 1547–1555 (2014).

51. Gudmundsson, J. et al. Common sequence variants on 2p15 and Xp11.22 confer susceptibility to prostate cancer. Nat Genet 40, 281–283 (2008).

52. Mamidi, T. K. K., Wu, J. & Hicks, C. Integrating germline and somatic variation information using genomic data for the discovery of biomarkers in prostate cancer. BMC Cancer 19, 229 (2019).

53. Hurwitz, L. M., Dogbe, N., Barry, K. H., Koutros, S. & Berndt, S. I. Obesity and prostate cancer screening, incidence, and mortality in the Prostate, Lung, Colorectal, and Ovarian Cancer Screening Trial. JNCI: Journal of the National Cancer Institute djad113 (2023) doi:10.1093/jnci/djad113.

54. Perez-Cornago, A., Dunneram, Y., Watts, E. L., Key, T. J. & Travis, R. C. Adiposity and risk of prostate cancer death: a prospective analysis in UK Biobank and meta-analysis of published studies. 2021.10.05.21264556 https://www.medrxiv.org/content/10.1101/2021.10.05.21264556v1 (2021) doi:10.1101/2021.10.05.21264556.

55. Hockla, A. et al. PRSS3/Mesotrypsin is a therapeutic target for metastatic prostate cancer. Molecular cancer research: MCR 10, 1555 (2012).

56. Hagenlocher, C. et al. ER stress-induced cell death proceeds independently of the TRAIL-R2 signaling axis in pancreatic β cells. Cell Death Discov. 8, 1–9 (2022).

57. Hernandez-Cueto, A. et al. Death receptor 5 expression is inversely correlated with prostate cancer progression. Molecular Medicine Reports 10, 2279–2286 (2014).

58. Subbiah, V. et al. Preclinical Characterization and Phase I Trial Results of INBRX-109, A Third-Generation, Recombinant, Humanized, Death Receptor 5 Agonist Antibody, in Chondrosarcoma. Clin Cancer Res 29, 2988–3003 (2023).

59. Forero-Torres, A. et al. TBCRC 019: An open label, randomized, phase II trial of nanoparticle albumin-bound paclitaxel (nab-PAC or Abraxane®) with or without the anti-death receptor 5 (DR5) monoclonal antibody tigatuzumab in patients with metastatic triple negative breast cancer. Clin Cancer Res 21, 2722–2729 (2015).

60. Reed, M. R. & Woolley, L. T. Uses of tranexamic acid. Continuing Education in Anaesthesia Critical Care & Pain 15, 32–37 (2015).

61. Didiasova, M., Wujak, L., Wygrecka, M. & Zakrzewicz, D. From Plasminogen to Plasmin: Role of Plasminogen Receptors in Human Cancer. Int J Mol Sci 15, 21229–21252 (2014).

62. Capello, M., Ferri-Borgogno, S., Cappello, P. & Novelli, F. α-Enolase: a promising therapeutic and diagnostic tumor target. FEBS J 278, 1064–1074 (2011).

63. Kwaan, H. C. & McMahon, B. The role of plasminogen-plasmin system in cancer. Cancer Treat Res 148, 43–66 (2009).

64. University of Aarhus. Perioperative Treatment With Tranexamic Acid in Melanoma; Prognostic and Treatment-related Impact of the Plasminogen-plasmin Pathway. https://clinicaltrials.gov/study/NCT05899465 (2023).

65. Northwestern University. Phase I/II Trial of In Vivo Angiostatin Generation With Tissue Plasminogen Activator (tPA) and Captopril in Patients With Progressive, Metastatic Cancer. https://clinicaltrials.gov/study/NCT00086723 (2012).

66. El-Akawi, Z. J., Abu-awad, A. M. & Khouri, N. A. Alpha-1 Antitrypsin Blood Levels as Indicator for the Efficacy of Cancer Treatment. World J Oncol 4, 83–86 (2013).

67. Liao, H.-Y., Da, C.-M., Liao, B. & Zhang, H.-H. Roles of matrix metalloproteinase-7 (MMP-7) in cancer. Clinical Biochemistry 92, 9–18 (2021).

68. Rosenbaum, E. et al. Marimastat in the treatment of patients with biochemically relapsed prostate cancer: a prospective randomized, double-blind, phase I/II trial. Clin Cancer Res 11, 4437–4443 (2005).

69. Bull, S. C. & Doig, A. J. Properties of Protein Drug Target Classes. PLoS One 10, e0117955 (2015).

